# Population Optimally Immunized after Accounting for Type-Specific COVID-19 Vaccine Waning Intervals: State-Level Prevalence and Trends

**DOI:** 10.1101/2021.12.09.21267295

**Authors:** Elizabeth B. Pathak, Jason L. Salemi

## Abstract

**BACKGROUND:** COVID-19 vaccines exhibit real-world waning effectiveness against SARS-CoV-2 infection within the first 3-6 months after a completed series. Consequently, the main metric tracked by the CDC (percent “fully vaccinated,” with no adjustment for booster status) has become insufficiently informative.

**METHODS:** We analyzed CDC daily vaccination data to quantify COVID-19 immunization status for 4 mutually-exclusive groups: (1) *not immunized;* (2) *partially immunized (*people who received the 1^st^ dose of a 2-dose series); (3) *immunized with waning immunity* (previously immunized people whose booster dose is overdue); and (4) *optimally immunized* (people who: (a) received the Janssen vaccine <2 months ago or completed an mRNA vaccine series <6 months ago, or (b) received the Janssen vaccine >2 months ago or completed an mRNA vaccine series >6 months ago and received a booster dose.)

**RESULTS:** The proportion of the total US population who were optimally immunized against COVID-19 fell from a high of 45.3% on July 17 to 29.4% on November 30. During November, the majority of states experienced a worsening trend in the percent of the total population who were overdue for a booster dose, including the 4 largest states, with percentage point increases of 3.5 in New York, 3.4 in California, 2.3 in Texas and 1.7 in Florida.

**CONCLUSIONS:** Our proposed classification scheme accounts for type-specific vaccine waning intervals, provides an accurate assessment of progress toward national immunization goals, and reveals the urgent need for additional public health mitigation strategies to successfully combat the COVID-19 pandemic in the United States.

## BACKGROUND

COVID-19 vaccines are highly efficacious in clinical trials,^1^ but exhibit real-world waning effectiveness against SARS-CoV-2 infection within the first 3-6 months after a completed series.^2–4^ National governments have responded to the evolving science on COVID-19 vaccine effectiveness with varying degrees of urgency. Israel began an aggressive campaign to administer 3^rd^ doses after 5 months to all adults on 30-Jul 2021.^3^ The US approved 3^rd^ doses for the immunocompromised after 6 months on 13-Aug.^5^ On 23-Sept, a Pfizer booster after 6 months was approved for the elderly, long-term care patients, medically high-risk persons aged 50-64 years, and occupationally high-risk adults 18-64 years old – but only for those who initially received the Pfizer vaccine.^5^ On 21-Oct, any booster dose was approved for any brand of initial vaccine series, for the 23-Sept risk categories, and for any Janssen vaccine recipient after 2 months.^5^ On 19-Nov, the CDC approved any booster dose 6 months after an mRNA vaccine (i.e. Pfizer or Moderna) for all adults 18+ years old.^6^

In the U.S., current surveillance of population vaccination status focuses on tracking the population who are “fully vaccinated,” defined by the CDC as those persons who have completed a vaccine series, no matter how many months in the past. This definition was unproblematic until the summer of 2021 when the first scientific reports of waning effectiveness were released from Israel^3^ and the U.S.^2^ Now, amid accumulating evidence of real-world waning effectiveness^4,7^ and growing numbers of “breakthrough” infections, including those resulting in hospitalization and death, there is an urgent need for a new approach. In this study, we define a new 4-category classification of COVID-19 immunization status which incorporates the exact date of completed immunization series, brand of vaccine received, and date of booster receipt to quantify the proportion of the population with waning immunity. We report the population prevalences for each mutually exclusive group, by state, on 30-Nov 2021, and examine one-month trends in state immunization status from 1-Nov to 30-Nov.

## METHODS

As illustrated in Figure 1, we define 4 mutually exclusive categories of COVID-19 vaccination status, and propose new nomenclature to replace (or complement) the “fully vaccinated” language which is currently emphasized by the CDC and widely used. We use the term “immunized” rather than “vaccinated” to better reflect the functional purpose of vaccine doses, and to sidestep a word that has become highly politicized during the pandemic. The ***not immunized*** population have never received a single COVID-19 vaccine dose. The ***partially immunized*** population have received the 1^st^ dose of a 2-dose regimen (i.e. either Moderna or Pfizer vaccine). The ***immunized with waning immunity (booster dose overdue)*** population are those who are ≥2 months past the Janssen vaccine or ≥6 months past a 2-dose completed mRNA vaccine series, and have not yet received a booster dose. The ***optimally immunized*** population are those who meet any of the following criteria: (1) received the Janssen vaccine within the past 2 months, (2) received the Janssen vaccine more than 2 months ago and received a booster dose, (3) completed an mRNA vaccine series within the past 6 months, (4) completed an mRNA vaccine series more than 6 months ago and received a booster dose.

**Figure 1.**
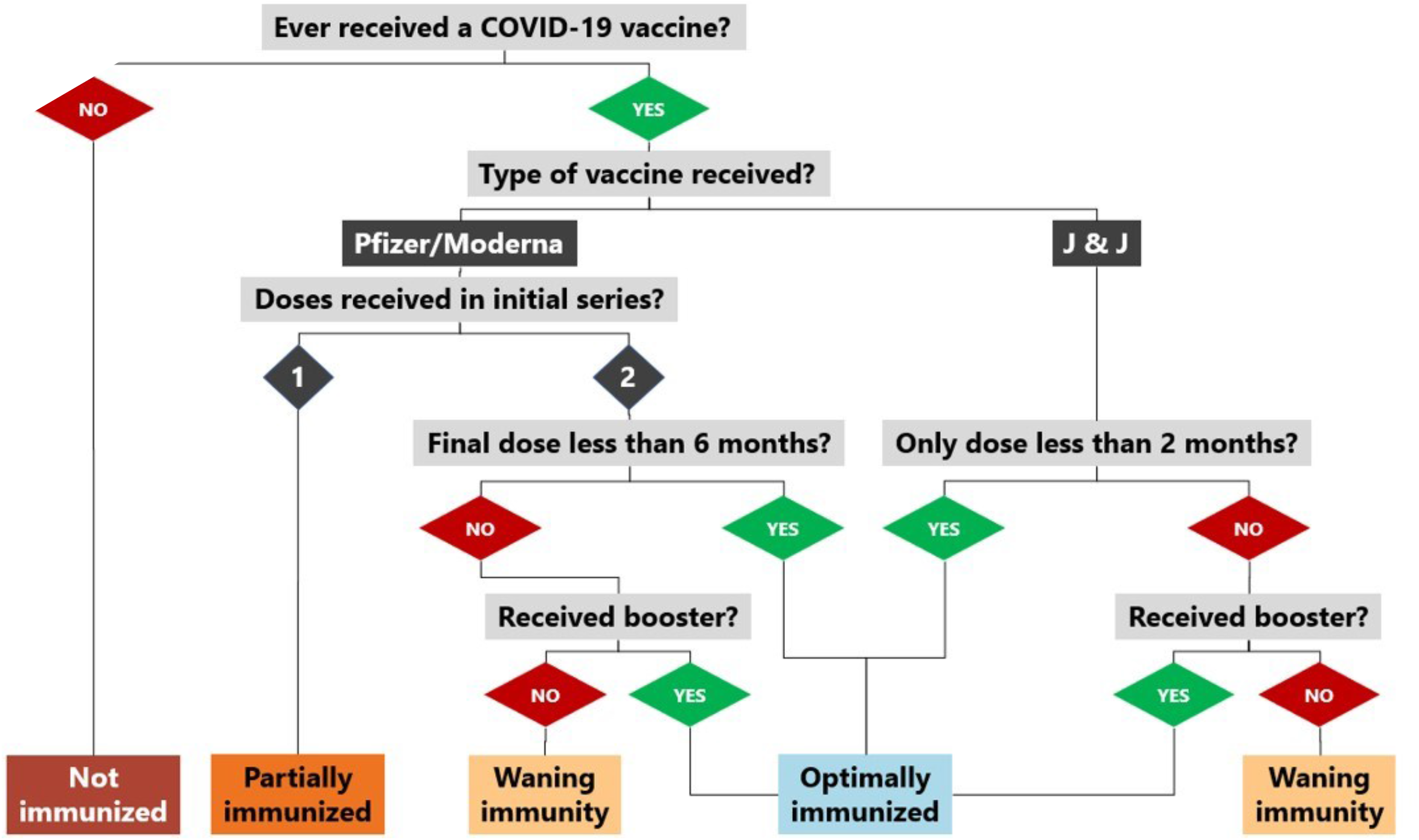
Decision Tree for Assigning COVID-19 Immunization Status Under New Proposed Classification

Vaccination data are reported by every state health department to the CDC^8^ on a daily basis. The data are reported in aggregate (not individual person) format, based on the following tabulation variables: (a) age group (all ages, 5+, 12+, 18+, 65+), (b) brand of vaccine (Janssen, Pfizer, Moderna, unknown 2-dose series), (c) date of 1^st^ dose (for 2-dose series), (d) date of series completion (either date of single Janssen dose or date of second Pfizer/Moderna dose). We applied simple arithmetic formulas to these aggregated data to assign the total population (2020 census population) of each state to one of the 4 mutually exclusive immunization categories for each day. For November 30, 2021, the calculations are as follows:

***Not immunized*** = total 2020 population subtracted by the cumulative number of people who had ever received at least one COVID-19 vaccine dose as of the index date (11/30/2021).

***Partially immunized*** = cumulative number of people who had received *only* the first dose of Pfizer/Moderna/unknown 2-dose series as of the index date (11/30/2021).

***Immunized with waning immunity*** = cumulative number of people who had a completed series 180 days prior to the index date (6/3/2021) for Pfizer/Moderna/unknown or 60 days earlier (10/1/2021) for Janssen. These are the people presumed to have waning immunity without a booster dose. We then subtracted the cumulative number of people who had received a booster dose as of the index date (11/30/2021).

***Optimally immunized*** = we summed three different groups: 1) cumulative number of people who completed a 2-dose Pfizer/Moderna/unknown series within the past 180 days (6/4/2021 to 11/30/2021); 2) cumulative number of people who received a Janssen dose within the past 60 days (10/2/2021 to 11/30/2021); and 3) the cumulative number of people with a booster dose as of the index date (11/30/2021).

Using these methods, we estimated (1) the percent of the total US 2020 population who fell into each immunization category on 30-Nov 2021 and (2) the one-month change (1-Nov to 30-Nov) in the percent of the total population who were overdue for a booster dose (immunized with waning immunity).

## RESULTS

The earliest COVID-19 vaccine recipients received their first doses of the Pfizer vaccine in December 2020. As can be seen in Figure 2, the proportion of the total population who were “fully vaccinated” and *optimally immunized* was nearly identical from December 2020 until June 2021. In June, our algorithm begins to reclassify individuals into the *immunized with waning immunity* group. At this point the curves begin to diverge. The proportion *optimally immunized* continued to increase for another month, because the number of individuals newly completing their vaccination series in June continued to outnumber the group who were reclassified as waning on each day. However, in July, the percent of the total population who were optimally optimized began to decline, and this decline continued for several months. At the end of October, when the FDA approved booster doses for a significant minority of the adult population, there was a slight increase in the percent optimally immunized immediately after FDA approval. Thereafter the trend plateaued, with a slight downward trend. At the very end of November, a slight upward trend (improvement) begins, after FDA approval of boosters for 18+ year olds and the 19-Nov CDC recommendation that all adults receive a booster regardless of age or medical conditions.

**Figure 2.**
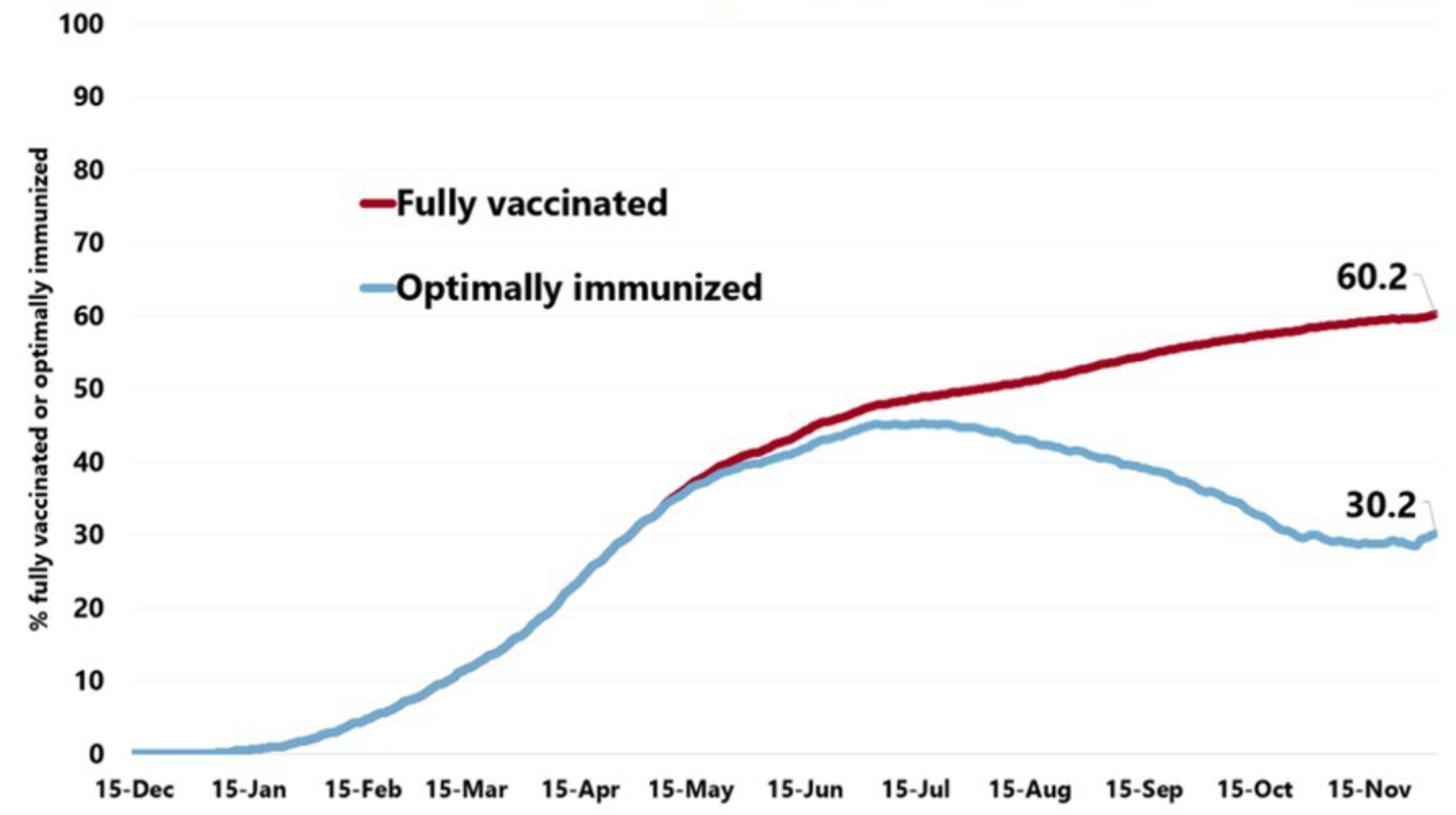
Comparison of the United States Temporal Trends in Percent of Total Population Fully Vaccinated vs. Percent Optimally Immunized

By 30-Nov 2021, the proportion of the US population who were optimally immunized had fallen to 29.4% (n=96.9 million) (Figure 3) from a high level of 45.3% on July 17. There were 96.7 million people (29.3%) who were not immunized on 30-Nov (including age-ineligible young children), 36.0 million (10.9%) who were partially immunized, and 99.9 million (30.3%) who were immunized with waning immunity. Only 6 states had more than one-third of their total population optimally immunized on 30-Nov (VT 40.1%, RI 38.0%, IL 35.3%, ME 34.6%, MD 34.3%, CT 33.6%) (Figure 3).

**Figure 3.**
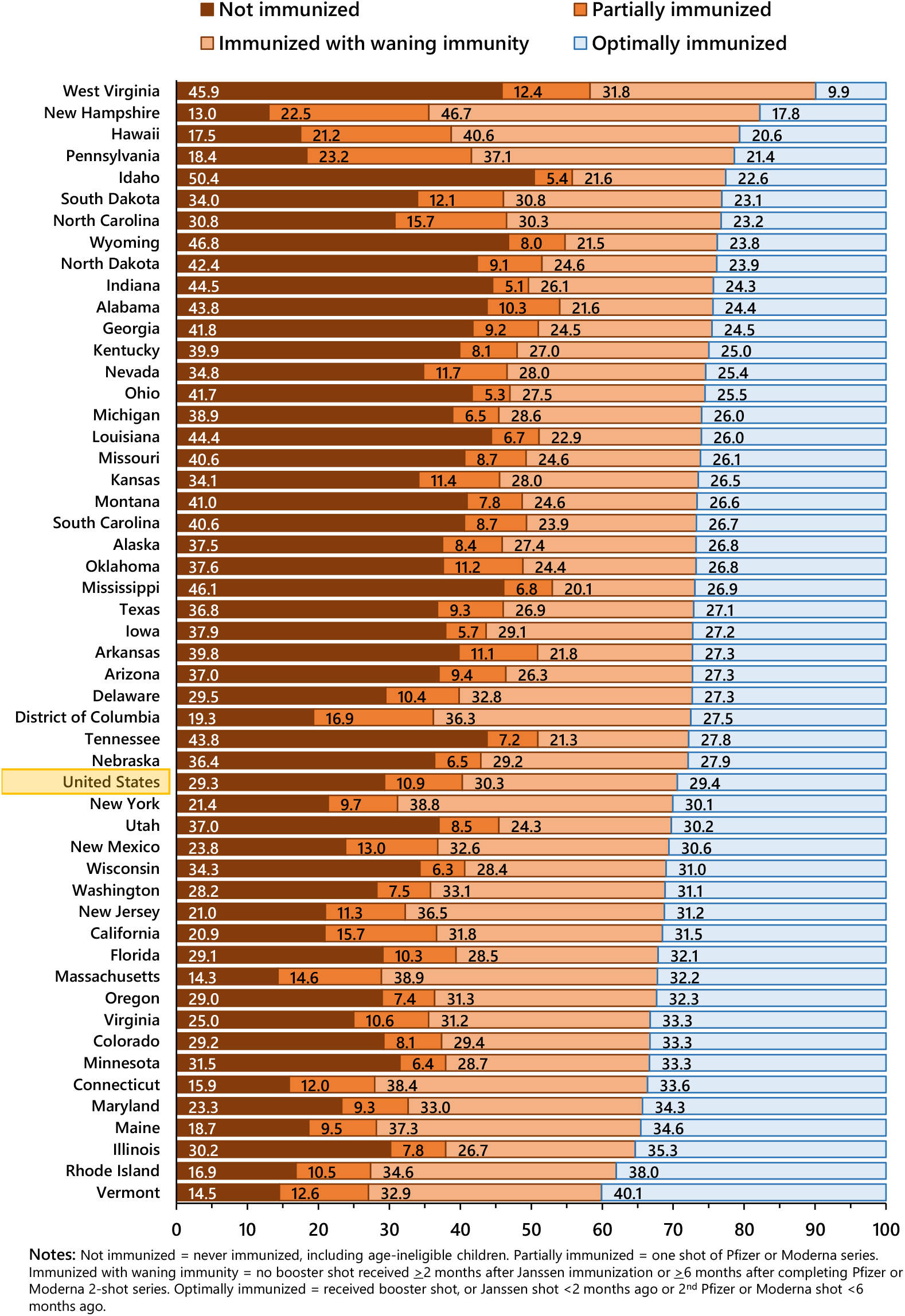
COVID-19 Immunization Status for States on November 30, 2021 Percent of 2020 Population (All Ages) **Notes:** Not immunized = never immunized, including age-ineligible children. Partially immunized = one shot of Pfizer or Moderna series. Immunized with waning immunity = no booster shot received ≥2 months after Janssen immunization or ≥6 months after completing Pfizer or Moderna 2-shot series. Optimally immunized = received booster shot, or Janssen shot <2 months ago or 2^nd^ Pfizer or Moderna shot <6 months ago.

From 1-Nov to 30-Nov, the proportion of the U.S. population who were overdue for a booster dose (immunized with waning immunity) increased slightly from 28.5% to 30.3%. Among states, there were 19 states which experienced improvement (i.e. declining percent who needed a booster dose), while the majority of states experienced a worsening trend (Figure 4). The percent of the total population who were overdue for a booster dose increased (worsened) in the 4 largest states, with a percentage point increase of 3.5 in New York, 3.4 in California, 2.3 in Texas and 1.7 in Florida.

**Figure 4.**
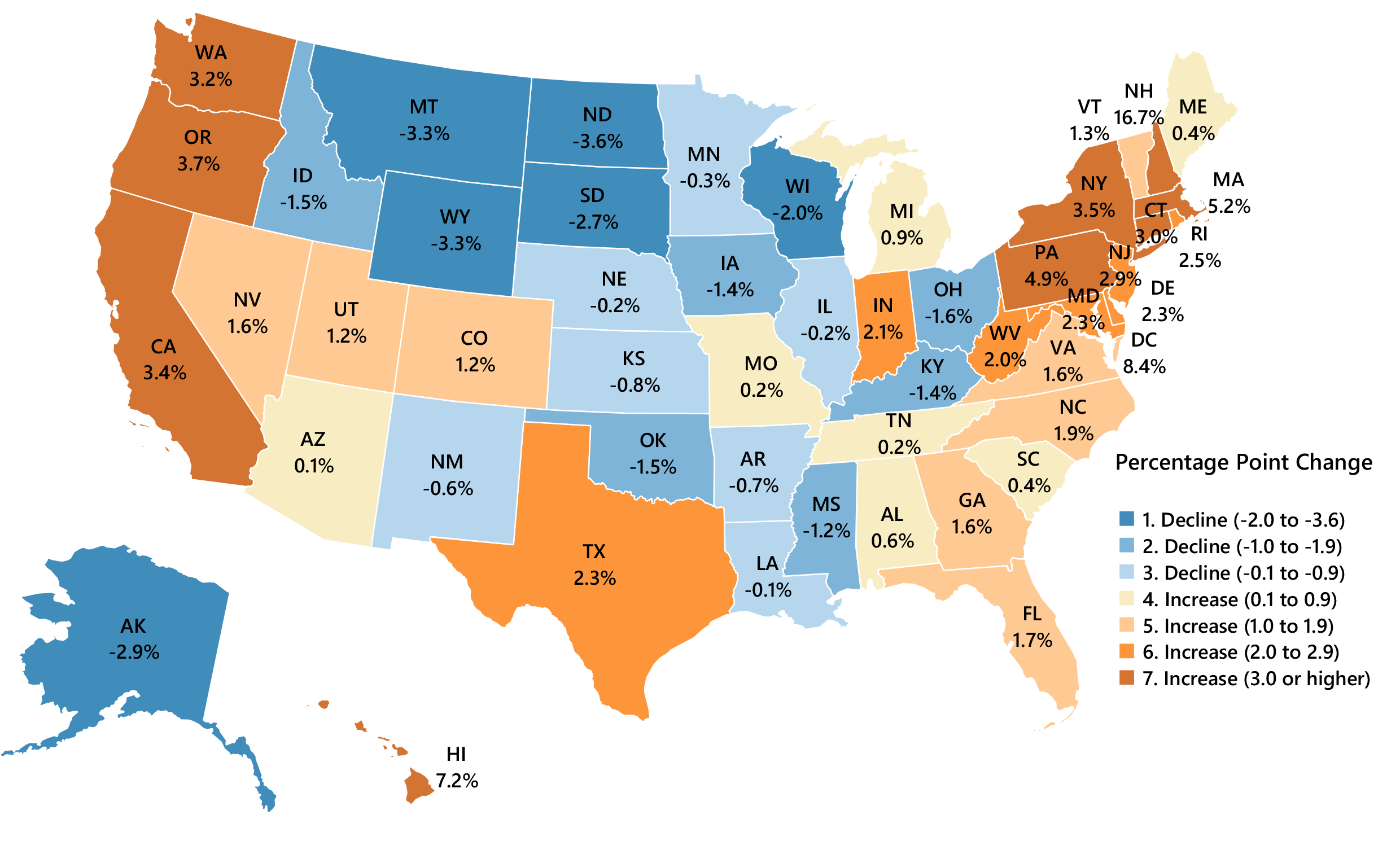
One-Month Change in the Percent of Total Population who were Overdue for a COVID-19 Vaccine Booster Dose: 1-Nov to 30-Nov 2021 *(Booster shot overdue = no booster shot ≥2 months after J&J vaccine or ≥6 months after Pfizer/Moderna 2nd dose)*

## DISCUSSION

The majority of the US population, in every state, was weakly or not immunized against COVID-19 immediately after the 2021 Thanksgiving holiday, based on the new immunization classification scheme proposed in our study. In contrast, according to the current CDC classification, the majority of the US population (∼60%) were “fully vaccinated,” with the concomitant connotation of being “fully protected” from COVID-19 at the end of November.^9^

Moreover, examination of one-month temporal trends revealed that several states which have previously been identified as “highly vaccinated states” (e.g. New England), along with the 4 largest states (California, Texas, Florida, and New York) all experienced adverse (increasing) trends from 1-Nov to 30-Nov in the percent of the total population who were overdue for a booster dose. We focused on the population overdue for a booster dose because from a pragmatic public health perspective, these individuals are “low-hanging fruit” who have already previously consented to being immunized against COVID-19.

In contrast, adults who have not yet consented to any COVID-19 vaccine may be a more challenging group with which to make progress. Historically, uptake of recommended vaccines in the general adult U.S. population has been far lower than the levels being targeted for COVID-19. Over a recent ten-year period, adult vaccine coverage rates for influenza, HPV, and Herpes zoster never exceeded 50%, according to the CDC.^10^ There remain immense challenges to adult vaccine uptake that must be addressed as efforts continue to reach, persuade, and vaccinate a high proportion of the US population.

There are several advantages to our proposed COVID-19 immunization surveillance categories and associated nomenclature. First, these categories are agnostic to personal history of COVID-19 infection and illness. Second, they utilize detailed, daily, publicly-available data on cumulative vaccination by type of vaccine to precisely reflect the population who were immunized but now have waning immunity on any given date. Third, a future change in the underlying definition of ***optimally immunized*** (e.g. if a 4^th^ booster dose becomes recommended in the future) can be implemented without the need for a change in terminology. Fourth, the replacement of the term *vaccinated* with the term *immunized* depoliticizes government agency recommendations and re-centers the positive physiological benefits of COVID-19 immunizations.

We have developed and pilot-tested a SAS algorithm to calculate the proportion of the population who fall into each immunization category each day, using the publicly available state-level vaccination dataset compiled by the CDC. To make these statistics freely available, we have updated our COVID-19 immunization web dashboard^11^ (part of The COVKID Project) in preparation for ongoing weekly updates with immunization status results for 5-11 year olds, 12-17 year olds, 18-64 year olds, 65+ year olds, and all ages combined.

Limitations of our study are as follows. Conceptually, the primary limitation of our method is that we assign groups of individuals to the *immunized with waning immunity* category in a dichotomous fashion, based on time frames that reflect population averages from empirical studies and regulatory time cutpoints (i.e. before or after 2 months for Janssen recepients, and before or after 6 months for Pfizer/Moderna recepients). While these time cutpoints are evidence-based and not arbitrary, they still cannot capture any of the individual variability in the timing of waning vaccine effectiveness, nor do they capture the continuous nature of the process of individual waning immunity at the physiological level. Several other nations have chosen to begin their administration of booster doses at 4 months or 5 months post-completed mRNA vaccine series, based on assessment of the exact same scientific evidence considered by the US FDA and US CDC. Consequently, by implementing a 6-month (for mRNA recipients) transition to the *immunized with waning immunity* category, instead of an algorithm that reflects possible waning immunity prior to that cutpoint, we take a conservative approach that may underestimate of the true percent of people with waning immunity. It then follows that our method may also overestimate the percent of the population optimally immunized. Simply put, as bad as things look, they may actually be worse.

Methodologically, the primary limitation of our approach is the underlying quality and accuracy of the vaccination data reported to the CDC by state health departments. For example, we and others have noted that in some states, the number of elders 65+ years old who have received at least one dose appears to be falsely elevated, for reasons unknown to us. A final limitation is that currently our algorithm assumes no vaccine waning among children <18 years old. If the FDA approves and the CDC recommends boosters for children in the future, we will update the algorithm to the extent permitted by the data available from CDC.

In conclusion, we recommend that the CDC update its COVID-19 data tracker to report number and percent of individuals who are *not immunized, partially immunized, immunized with waning immunity*, and *optimally immunized*, for the nation as a whole and for individual states/jurisdictions, with each stratified by age. These proposed surveillance data provide a more accurate assessment of progress toward national immunization goals and reveal the urgent need for additional public health mitigation strategies to successfully combat the COVID-19 pandemic in the United States.

## Data Availability

All data produced in the present study are available upon reasonable request to the authors

http://covid19florida.mystrikingly.com/

## Acknowledgement

The authors thank Dr. Amit P. Pathak for his suggestion of the term “optimally immunized.”

